# Modelling and forecasting a potential chikungunya outbreak in France, 2025

**DOI:** 10.1101/2025.08.12.25333506

**Authors:** Sandeep Tegar, Dominic P. Brass, Bethan V. Purse, Antoine Mignotte, Guillaume Lacour, Christina A. Cobbold, Steven M. White

## Abstract

Mainland France is experiencing an unprecedented number of autochthonous cases of chikungunya in multiple locations, primarily in southern France, likely linked to the recent outbreak in La Réunion and associated travel. Given the region’s suitable climate and the widespread and abundant presence of *Aedes albopictus*, the primary European mosquito vector of chikungunya, further cases are anticipated in the coming months. Here, we apply a state-of-the-art, climate-sensitive eco-epidemiological model to forecast chikungunya transmission dynamics at affected outbreak locations. With a 2–3-month lead time in predictions of peak transmission, the model identifies foci most likely to experience severe outbreaks (e.g., Salon-de-Provence, Bernis, Castries) this year and helps to inform prioritisation and public health impact.

## Introduction

Chikungunya virus (CHIKV) is a debilitating *Aedes*-borne alphavirus which is usually endemic in tropical climates [1]. In recent history (2010-2024), a small number of autochthonous (locally transmitted) cases of CHIKV have been reported in mainland France with *Aedes albopictus* (Asian tiger mosquitoes) implicated in transmission. These historic outbreaks have occurred in only two or three locations at most in a year, typically involving a low number of cases and initiated by travellers returning from endemic regions—including in 2025. However, in 2025, France is experiencing emergence of an unprecedented chikungunya outbreak across a wide spatial extent. Since 2010—the year the first two autochthonous chikungunya cases were reported—a total of 32 autochthonous cases were recorded between 2010 and 2024 [2]. As of 30 July 2025, 48 autochthonous cases have already been reported in 2025 alone [3]. This unusual emergence of CHIKV in France is most likely associated with the large number of imported cases—833 reported since 1 May 2025 [3]—following a recent outbreak in La Réunion (August 2024 to May 2025), which has affected approximately 51,558 individuals to date [4].

The recent CHIKV outbreaks in mainland France are expected to intensify in the coming months— assuming no vector control is implemented—driven by favourable breeding conditions for the primary mosquito vector, *Ae. albopictus*, its high competence for CHIKV transmission, and the warm summer season, which enhances mosquito activity. Although reported CHIKV case numbers per city are still low [3], this is typical for *Aedes*-borne arbovirus outbreaks in Europe at this time of year [5]. Historical CHIKV outbreaks show that case counts often rise sharply in August and September [6-9]. However, the severity of the outbreaks is likely to be variable, ranging from a few isolated cases to large numbers of cases with an associated high disease burden [1]. Determining the likely outbreak severity in different locations is critical for timely planning and targeting of public health measures such as vector control, public awareness campaigns and healthcare burden mitigation [10].

In this study, we apply a state-of-the-art climate-sensitive mechanistic model to forecast simultaneously the population dynamics of *Ae. albopictus* and the transmission dynamics of CHIKV by *Ae. albopictus* in France through the end of 2025. We aim to identify locations where CHIKV outbreaks are likely to be most severe in the absence of effective mitigation measures to help inform public health decision-making.

### Modelling the *Ae. albopictus* abundance and CHIKV transmission dynamics

Our model builds on a previously validated, environmentally driven, stage- and phenotype-structured modelling framework designed to study dengue virus transmission by *Ae. albopictus* in Europe [11]. We adapted this dengue model to CHIKV transmission to conduct location-specific outbreak forecasts for each site in France where autochthonous cases have been reported (Figure 1): La Crau (Var, Provence-Alpes-Côte d’Azur), Prades-le-Lez (Hérault, Occitanie), Salon-de-Provence (Bouches-du-Rhône, Provence-Alpes-Côte d’Azur), Grosseto-Prugna (Corse-du-Sud, Corse), Montoison (Drôme, Auvergne-Rhône-Alpes), Bernis (Gard, Occitanie), Lipsheim (Bas-Rhin, Grand Est), Claix (Isère, Auvergne-Rhône-Alpes), Fréjus (Var, Provence-Alpes-Côte d’Azur), Saint-Brès/Castries (Hérault, Occitanie), Toulon (Var, Provence-Alpes-Côte d’Azur), and Saint-Chamond (Loire, Auvergne-Rhône-Alpes).

**Figure 1.**
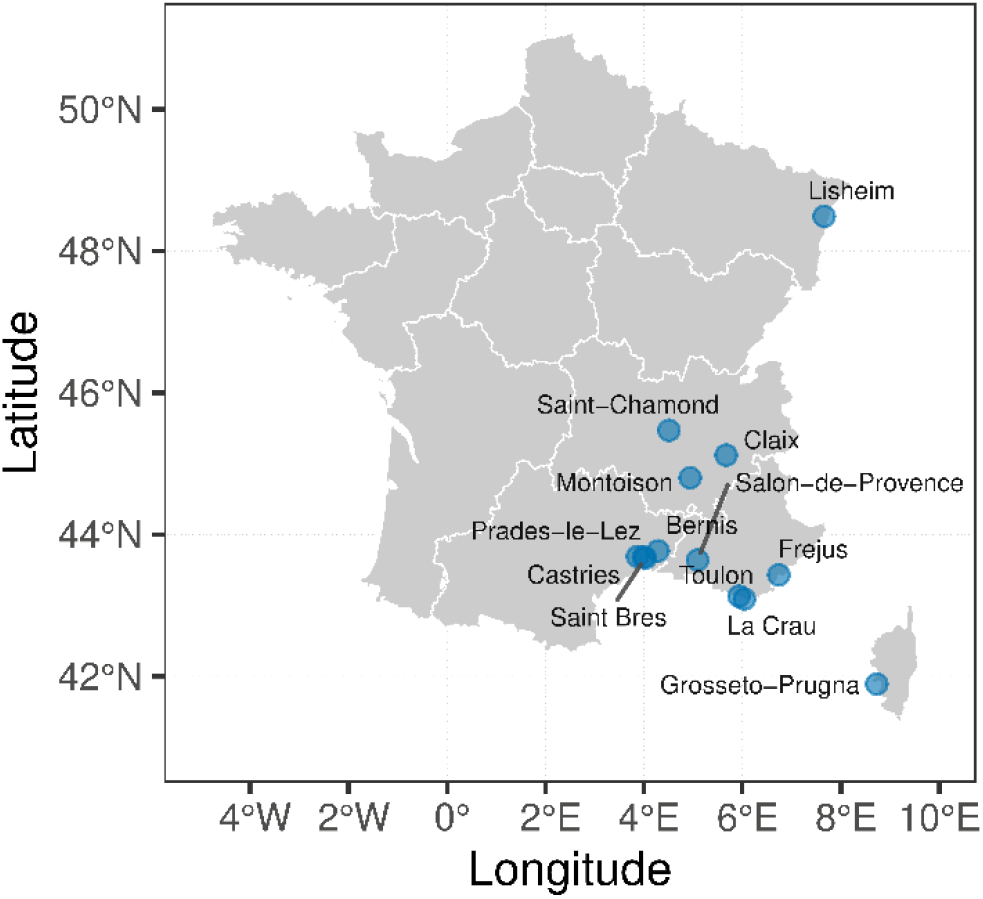
Locations in France where autochthonous cases of CHIKVD have been reported as of 16 July 2025 [3].

Our state-of-the-art dynamic model [11] characterises the transmission dynamics of CHIKV using a Susceptible-Exposed-Infected-Recovered (SEIR) epidemic framework, integrated with the environmentally driven, stage- and phenotype-structured population and trait dynamics of *Ae. albopictus* (Supplementary Figure S1). Environmentally driven trait dynamics represent the phenotypic plasticity of the invasive mosquito *Ae. albopictus* in novel environments, capturing in real time the eco-epidemiological traits relevant to mosquito-borne diseases, e.g., mosquito longevity, egg diapause and quiescence, development rate, stage-specific survival probabilities, biting rate, and the virus’s extrinsic incubation period (i.e., the time between a mosquito feeding on an infected human and becoming infectious) [11].

By modelling the intricate relationships between environmental conditions, mosquito traits, mosquito population dynamics, and virus traits, parameterised by extensive laboratory-derived data, the model can predict virus spread and mosquito population dynamics using remotely sensed human density and climate reanalysis data (Supplementary Figures S3-S16). The model has been extensively validated globally for seasonal *Ae. albopictus* population dynamics dengue outbreaks, showing high degrees of accuracy without backfitting the model [11]. Here, the model is parameterised for CHIKV transmission by incorporating the temperature-dependent relationships for *Ae. albopictus* and CHIKV vector competence and extrinsic incubation period and has been additionally validated against historic Italian CHIKV outbreaks.

To predict future CHIKV spread in France, location specific daily temperature, precipitation, and evaporation data were obtained from the ERA5-Land climate reanalysis dataset, accessed via the Copernicus Climate Data Store [12] covering the period from January 1, 2015, to June 13, 2025. For the remainder of 2025, climate conditions were approximated using historic daily values derived from environmental data spanning 2015 to 2024, resulting in the creation of 10 climate scenarios to forecast the future climate of 2025. Additionally, host human population density data were sourced from the gridded population dataset based on the 2021 Population and Housing Census, available through the Eurostat web portal [13]. The climate data have a spatial resolution of 0.1° × 0.1°, while the population data have a resolution of 1 km^2^. Since the model operates at a spatial resolution of 2 km × 2 km, both population density and climate data were resampled to a grid size of 4 km^2^. Since the data are preliminary, and without further information, we used the likely date of the index case as a proxy for the primary case, as reported in the weekly bulletin of the French national public-health agency [3], to initiate the outbreak in our model simulations.

### Forecasting the 2025 CHIKV outbreaks in France

For each location in France where autochthonous CHIKV cases were reported up to 16 July 2025 [3], we analysed the possible outbreak onset or take-off window (i.e., a time period that marks the interval starting from when the (simulated) daily number of new (autochthonous) cases first exceeds one and continues to grow, ultimately leading to the peak of the outbreak – a measure of the exponential phase of the outbreak), average peak incidence (i.e., average number of daily cases at the peak), and the final outbreak size (i.e., total cumulative number of cases at the end of season) (Figure 1).

Following the first reported symptomatic cases [3], we simulated the model with one symptomatic index case for all cities, except for Salon-de-Provence and Grosseto-Prugna, where two symptomatic index cases were used. We predict that, in the absence of any control or mitigation measures, large outbreaks could occur in Salon-de-Provence (2,102 cases (95% CI: 1,009.5–3,196.0)), Bernis (1,119.6 cases (95% I: 430.0–1,809.2)), Castries 413.8 (95% CI: 132.6-695.1), La Crau (220.0 cases (95% CI: 68.0–372.2)), Toulon (211.0 cases (95% CI: 76.0–384.2)), Prades-le-Lez (203.0 cases (95% CI: 60.6–344.9)), Saint-Brès (180.4 cases (95% CI: 52.4–301.3)), and Fréjus (135.6 cases (95% CI: 33.3– 213.2)) (Figure 2), with the potential for a moderately sized outbreak in Grosseto-Prugna (coastal) (31.7 cases (95% CI: 19.4–43.9)) and smaller outbreaks in the remaining locations (Table 1).

**Table 1.** Outbreak locations in France with geographic and demographic information, along with simulated estimates for the outbreak onset window, average peak incidence (i.e., average number of daily cases at peak), and final outbreak size (i.e., total cumulative number of cases at the end of epidemic). Values in parentheses in the projected total cases column represent the 95% confidence intervals (CIs), while the dates in parentheses in the outbreak onset window column indicate the range between the earliest and latest estimated onset dates.

| Region | Department | Commune | Symptom onset date for the first case | Lat/ Lon | Estimated population (per 4 km<sup>2</sup>) | Predicted window for outbreak take-off | Average peak incidence cases (Date) | Projected total case count |
| --- | --- | --- | --- | --- | --- | --- | --- | --- |
| Provence-Alpes-Côte d'Azur | Bouches-du-Rhône | Salon-de-Provence | 16/06/2025 | (43.64, 5.09) | 8,525 | 28/07/2025<br>(17/07/2025-09/08/2025) | 54.1<br>(20/09/2025) | 2,102.7<br>(1,009.5-3,196.0) |
| Occitanie | Gard | Bernis | 11/06/2025 | (43.77, 4.29) | 7,000 | 06/08/2025<br>(27/07/2025-17/08/2025) | 27.8<br>(24/09/2025) | 1,119.6<br>(430.0-1,809.2) |
| Occitanie | Hérault | Castries | 30/06/2025 | (43.68, 3.99) | 6,014 | 16/08/2025<br>(04/08/2025-25/08/2025) | 9.9<br>(24/09/2025) | 413.8<br>(132.6-695.1) |
| Provence-Alpes-Côte d'Azur | Var | La Crau | 02/06/2025 | (43.09, 6.04) | 10,637 | 31/08/2025<br>(13/08/2025-26/09/2025) | 4.3<br>(02/10/2025) | 220.0<br>(68.0-372.2) |
| Provence-Alpes-Côte d'Azur | Var | Toulon | 16/06/2025 | (43.13, 5.93) | 31,997 | 30/08/2025<br>(12/08/2025-17/09/2025) | 4.0<br>(28/09/2025) | 211.0<br>(76.0-384.2) |
| Occitanie | Hérault | Prades-le-Lez | 27/05/2025 | (43.69, 3.87) | 6,000 | 25/08/2025<br>(08/08/2025-09/09/2025) | 4.8<br>(20/09/2025) | 203.0<br>(60.6-344.9) |
| Occitanie | Hérault | Saint Bres | 30/06/2025 | (43.67, 4.03) | 4,094 | 26/08/2025<br>(10/08/2025-09/09/2025) | 4.2<br>(20/09/2025) | 179.7<br>(58.4-301.1) |
| Provence-Alpes-Côte d'Azur | Var | Fréjus | 01/07/2025 | (43.43, 6.74) | 12,975 | 04/09/2025<br>(19/08/2025-19/09/2025) | 2.7<br>(26/09/2025) | 135.6<br>(33.3-213.2) |
| Corse | Corse du Sud | Grosseto-Prugna (coastal) | 19/06/2025 | (41.89, 8.73) | 3,173 | NA | 0.52<br>(17/09/2025) | 31.7<br>(19.4-43.9) |
| Auvergne-Rhône-Alpes | Drôme | Montoisson | 13/06/2025 | (44.80, 4.94) | 1,460 | NA | 0.1<br>(11/09/2025) | 4.4<br>(1.3-7.4) |
| Auvergne-Rhône-Alpes | Loire | Saint-Chamond | 01/07/2025 | (45.47, 4.51) | 20,000 | NA | 0.1<br>(06/09/2025) | 3.0<br>(1.2-5.0) |
| Grand Est | Bas-Rhin | Lipsheim | 26/06/2025 | (48.49, 7.66) | 2,643 | NA | 0.04<br>(08/09/2025) | 2.0<br>(1.2-3.0) |
| Auvergne-Rhône-Alpes | Isère | Claix | 01/07/2025 | (45.12, 5.67) | 6,000 | NA | 0.01<br>(13/08/2025) | 0.5<br>(0.2-0.8) |

**Figure 2.**
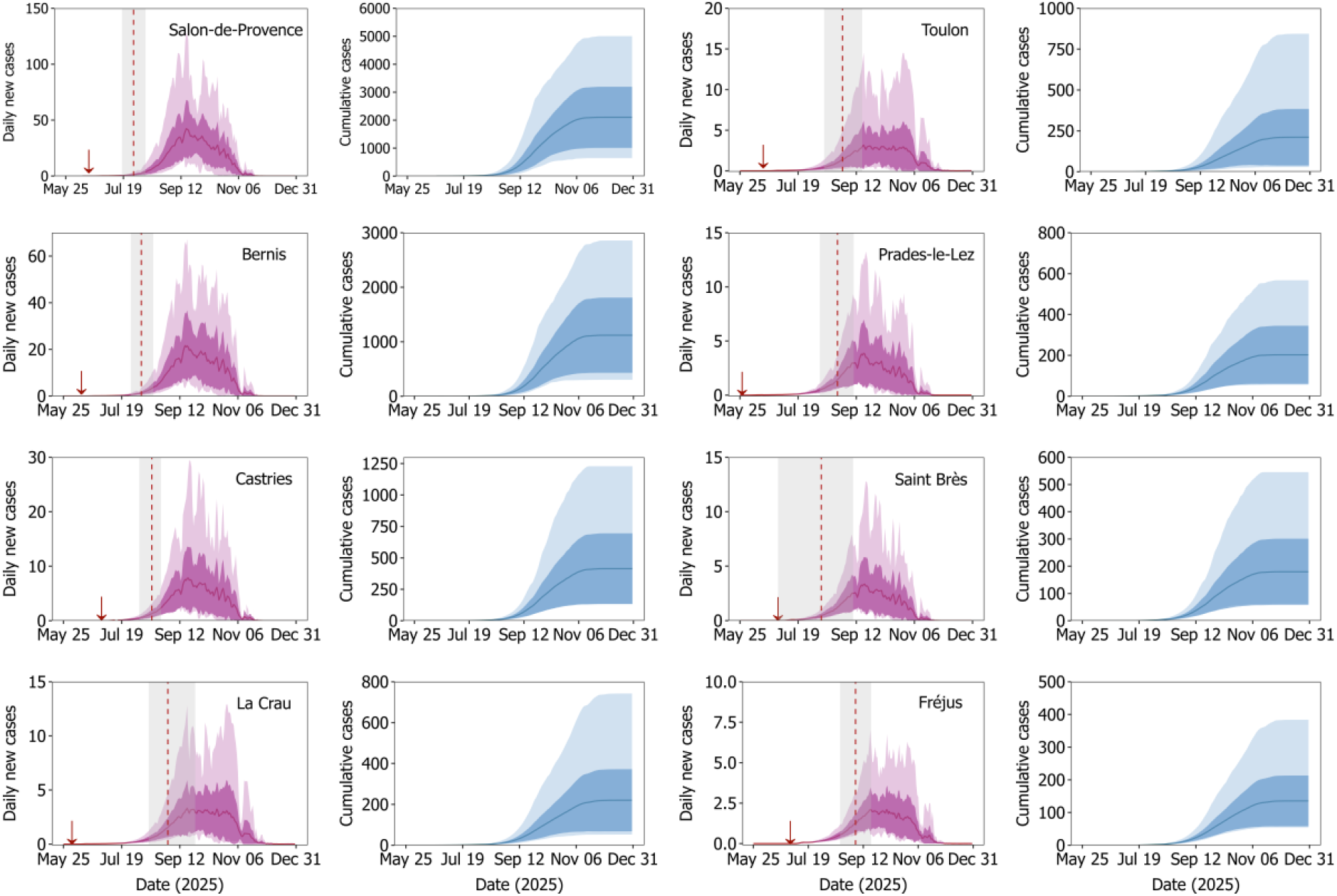
CHIKV transmission dynamics for locations in France where large outbreaks could occur. For each location shown in the panels above, the red plots represent the daily number of cases. The vertical red dashed lines indicate the estimated mean onset date of the outbreak, while the surrounding grey bands represent the range between the earliest and latest estimated onset dates. The blue plots show the cumulative number of cases. In all plots, the solid lines represent the mean values, the dark ribbons indicate the 95% confidence intervals (CIs), and the light-coloured bands correspond to the absolute maximum and minimum values. The introduction dates, indicated by red downward arrows in the plots, correspond to the first reported symptomatic case in each location (Table 1). The variability in the projections reflects stochasticity in climate variables—temperature, precipitation, and evaporation—for the remainder of the year 2025. See Supplementary Figure S2 for the locations where the CHIKV outbreak severities are predicted to be low.

We predict that the outbreak could begin between the second and third weeks of August 2025 in all studied locations with a relatively high potential for large outbreaks, with Salon-de-Provence, Bernis, and Castries possibly experiencing onset as early as 28 July to 16 August 2025 (Table 1). In contrast, La Crau, Toulon, and Fréjus in the Var department may experience onset as late as 25 August to 4 September 2025.

The peak incidence is predicted to occur between late September and early October 2025 (20/09/2025– 02/10/2025) across all outbreak locations with the potential for larger outbreak sizes. The maximum peak incidence is estimated to reach approximately 54 cases per day on average in Salon-de-Provence, surpassing the projected maximum peak incidence of 28 daily cases in Bernis and 10 daily cases in Castries (Table 1).

Although our results suggest a robust prediction in total case numbers, there is substantial uncertainty in our predictions owing to variation in weather conditions. European arbovirus transmission is highly sensitive to temperature variation, with warmer weather associated with increased mosquito fecundity, shorter generation times and faster transmission [11]. Furthermore, sharp changes in temperature induced by extreme precipitation can affect mosquito biting and therefore provide a climate-induced break in transmission leading to a reduction in cases [14].

In addition, for major outbreak locations where cases are already relatively high (e.g., Salon-de-Provence, Castries, and Grosseto-Prugna (coastal)) [3, 15], we compared the reported cases with model predictions for these locations to estimate the primary case date. To achieve this, we vary the primary case date in the model and minimise the RMSE between predicted cumulative cases and the data utilising the most recent reported cases notified up to 30 July 2025 [15], matching the comparison dates the to the timing of the reports.

Our model predictions show broad agreement with this initial phase of outbreak in these cities when adjusting for the more likely primary case date, each showing an increase in cases (Figure 3). Our model predicts that the primary case dates to be 12/06/2025 for Grosseto-Prugna (coastal) compared to the earliest known index case of 19/06/2025, 15/06/2025 for Castries compared to the earliest known index case of 30/06/2025, and 29/05/2025 for Salon-de-Provence compared to the earliest known index case of 16/06/2025. It should be noted that these estimates are based on small amounts of data that may be latterly revised. Additional case data over the course of the outbreaks and case tracing are highly likely to improve on our estimates of these primary case dates. However, these estimates provide a useful early indication and only have a small effect on the predicted number of cases (Figure 3).

**Figure 3.**
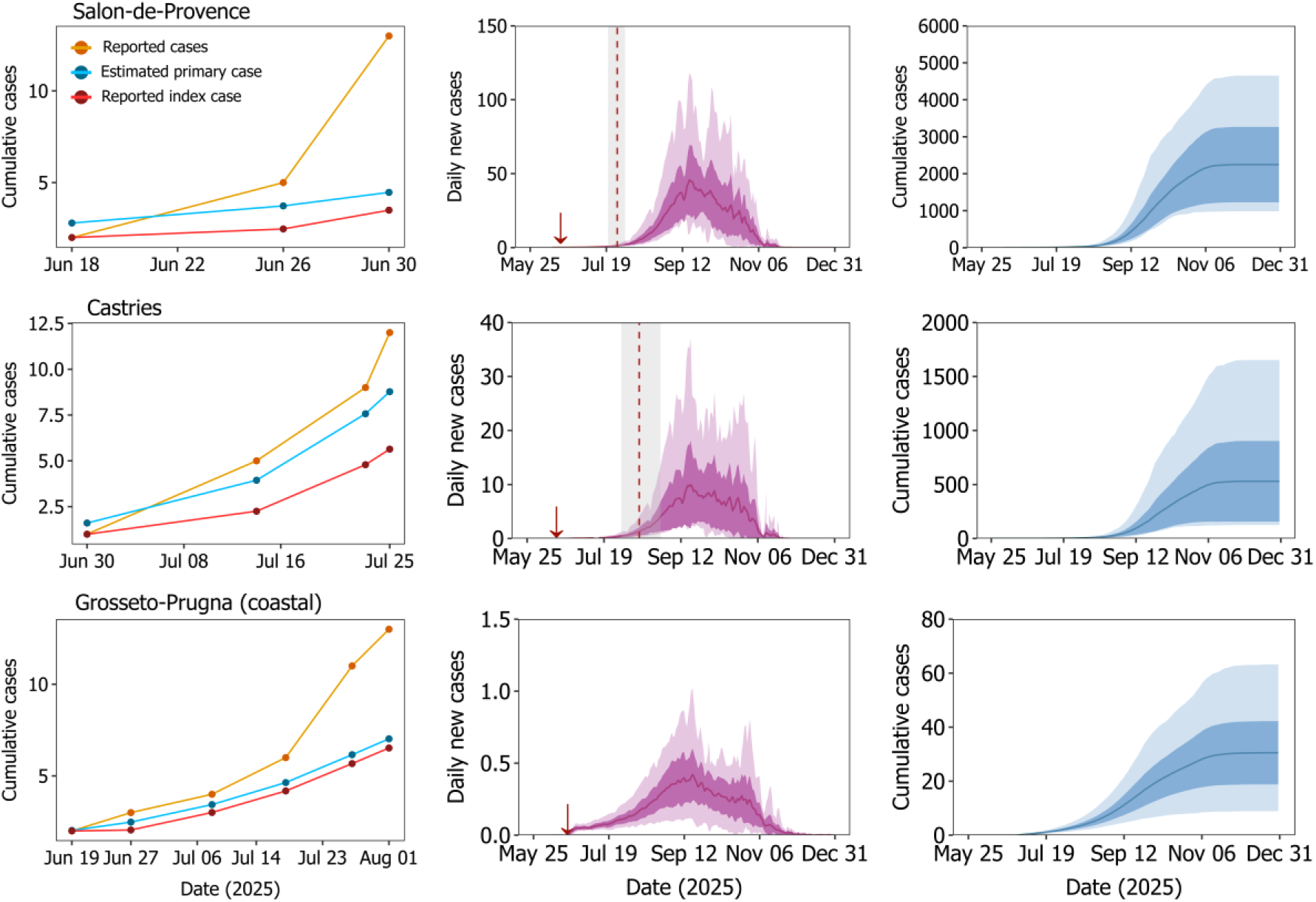
Early phase of chikungunya transmission in Salon-de-Provence, Castries, and Grosseto-Prugna (coastal). The leftmost panel shows reported data up to 01/08/2025 in orange [15]. The blue and red lines represent the model outputs generated using introduction dates for the estimated primary cases and the reported index cases (index case dates listed in Table 1). The estimated introduction dates for the primary cases are 29/05/2025 for Salon-de-Provence, 15/06/2025 for Castries, and 12/06/2025 for Grosseto-Prugna (coastal). For each location shown in the panels above, the red plots (middle column) represent the daily number of cases. The vertical red dashed lines indicate the estimated mean onset date of the outbreak, while the surrounding grey bands represent the range between the earliest and latest estimated onset dates. The blue plots (right column) show the cumulative number of cases. In all plots, the solid lines represent the mean values, the dark ribbons indicate the 95% confidence intervals (CIs), and the light-coloured bands correspond to the absolute maximum and minimum values. The estimated introduction dates for the primary cases indicated by red downward arrows in the plots.

## Discussion

Previous years autochthonous CHIKV outbreaks in France were limited to a maximum of three locations (predominantly Occitanie and Provence-Alpes-Côte d’Azur) [2]. In contrast, the situation in 2025 is unprecedented, with an ongoing outbreak currently affecting more than 12 administrative units across the southern region of the country [3], with autochthonous transmission in Nouvelle-Aquitaine, Auvergne-Rhône-Alpes, and Grand Est regions, with markedly different climates and mosquito vector densities. Although autochthonous chikungunya case data indicate that only a small number of cases have been reported so far, this likely reflects an early-stage trend. A significant escalation may occur during August–September 2025, as observed in previous *Ae. albopictus*-borne chikungunya virus (CHIKV) outbreaks in Europe [6-9]. In this context, our CHIKV forecast study, based on a climate-sensitive eco-epidemiological dynamic model, quantifies the likelihood of such an escalation at each affected location across all 12 administrative units in France (Table 1). The model also successfully predicts the early signs of CHIKV escalation in three distinct foci that are showing substantial increases in cases (Figure 3), predicting likely earlier primary index cases compared to the currently known earliest index cases.

The climate-sensitive predictions of *Ae. albopictus* population dynamics and CHIKV transmission 2–3 months in advance of transmission peak—particularly during the period when major escalation in cases is likely—can significantly enhance the timeliness and effectiveness of public health responses to the outbreak. Our CHIKV forecasting model provides estimates of the overall number of cases for the current season, as well as the intensity and timing of peak incidence, thus generating a “wake-up call” and reliable guidance regarding the potential scale of transmission. Such information can be utilised for public health decision-making on when and where to initiate control efforts, thereby enhancing surveillance, early diagnostic testing, and healthcare preparedness that may not be currently ready [16]. For instance, predictions regarding the timing of peak incidence—such as an estimated ∼54 daily new CHIKV cases in mid-September 2025 in Salon-de-Provence (Table 1)—can inform location-specific policy decisions on the required intensity and frequency of mosquito control programs, the need to scale up diagnostic and seroprevalence efforts, and strategies to interrupt the transmission cycle. Forecasts may be particularly valuable operationally prior to outbreak seasons, if the model is extrapolated across whole regions that are vulnerable to arrival of viral people through travel, for different seasonal timings of introduction.

We acknowledge that many of the predicted outbreak sizes may not be realised. This is most likely due to existing and active mitigation and control measures in the locations reporting CHIKV cases [3], which are likely to reduce the reduce the total number of cases by the end of the year or halt the outbreak altogether especially if the outbreak has been detected quickly and vector control methods are effective. Furthermore, the scale of the total number of cases is likely to be uncertain due to the effect of future weather on *Ae. albopictus* population dynamics. However, the predicted intensity and timing of peak incidence serve as reliable inputs for designing policy decisions in response to the larger outbreaks identified in our study—e.g., in Salon-de-Provence, Bernis, Castries, La Crau, Toulon, Prades-le-Lez, Saint-Brès, and Fréjus. Worryingly, our predictions indicate that Salon-de-Provence is likely to experience the largest CHIKV outbreak (Figure 2), which aligns with the reported and currently available data (Figure 3) [3], suggesting that either there is a delay in observing the effects of mitigation on CHIKV cases (which is highly likely [14, 17]) or that mitigation needs to be intensified.

Small scale CHIKV outbreaks have been observed in mainland France since 2010, with a single case reported in Paris in 2024 [2]. However, larger outbreaks have been observed in Italy with 330 cases in 2007 in Emilia Romagna and 270 cases in 2017 in Lazio [2], and more recently, over 50,000 cases in La Réunion [4]. The ongoing CHIKV outbreaks in mainland France shares similar environmental conditions (early summer importation of virus, favourable climate, established *Ae. albopictus* populations, and higher attack rates in immunologically naïve populations [18]) with previous outbreaks in Italy, suggesting a possible risk of larger outbreaks in mainland France compared to previous years, as forecasted by our model in some locations (Table 1). The large spatial extent of the 2025 outbreaks is more likely attributable to the importation pressure due to the ongoing La Réunion chikungunya outbreak and high degrees of travel between the island and mainland France.

In general, due to the establishment of *Ae. albopictus* and the extended summer season, the southern region of France is expected to experience a higher number of cases of *Ae. albopictus*-borne diseases. However, even within southern France, not all locations exhibit equal vulnerability to CHIKV transmission. For instance, the peak incidence for Toulon and Salon-de-Provence show striking contrasts (Table 1). Although Toulon is a larger urban area situated further south, the predicted outbreak potential for CHIKV is substantially greater in Salon-de-Provence (Figure 2). These differences are largely attributable to climatic variations: Toulon exhibits a Mediterranean coastal climate, whereas Salon-de-Provence experiences a Mediterranean climate influenced by semi-inland conditions with more pronounced summer temperature and rainfall (Supplementary Figure S17). Such climatic differences impact *Ae. albopictus* abundance and the timing of its annual activity cycle, which in turn affects the vector-to-human density ratio—a critical determinant of CHIKV transmission dynamics. The smaller outbreak size projected for Toulon corresponds with its relatively lower simulated *Ae. albopictus* abundance compared to Salon-de-Provence (Supplementary Figure S13, S15 and S17). In contrast, although Montoison shows *Ae. albopictus* abundance comparable to Salon-de-Provence, its lower human population density reduces the vector-to-human ratio, leading to a smaller predicted outbreak size (Table 1 and Supplementary Figure S2). This reasoning can also be extended to the observed differences in predicted outbreak sizes and take-off windows across all the studied locations (Table 1 and Supplementary Figures S3-S16).

Utilising minimal case data over time we have attempted to estimate the primary case date (Figure 3). In each scenario the model predicts an earlier primary case date compared to the index case dates reported by 1-2 weeks. In previous arbovirus outbreaks in Europe, including those caused by CHIKV, the time gap between the primary case and the index case can often near a month [7-9]. Although there is much uncertainty with our analysis due to a low number of data points, the results suggests that the primary cases have yet to be detected and that there may be additional cases that have yet to be reported (or some reported cases have an earlier symptom onset yet to be determined in future analysis). This is especially evident in the Salon-de-Provence outbreak where an additional 11 cases where identified within 1 week.

The speed of publishing information on CHIKV cases has enabled us to predict near real-time case numbers for the outbreak locations. We have been able to achieve this with minimal information, primarily the timing and location of the index case. Since the model has been extensively validated with historic dengue and chikungunya outbreak data (e.g., Brass et al. [11]), we may place some degree of confidence in the predictions, but strongly noting the uncertainty associated with climatic variation and mitigation measures could have on the courses of the outbreaks. With a regular flow of real-time case and environmental data, it should be possible to benchmark the effectiveness of public health mitigation strategies.

## Conclusion

A climate- and *Ae. albopictus* population-informed chikungunya forecast, as demonstrated in this study, can significantly improve the timeliness and effectiveness of public health responses to ongoing chikungunya outbreaks in France. By providing reliable inputs for decision-making, such forecasts support targeted mosquito-control measures, enhanced surveillance, and improved healthcare preparedness. Predictions of outbreak intensity and timing can serve as evidence to support the proactive deployment of emergency mosquito control measures and to ensure sufficient medical supplies and treatment capacity in the identified high-risk zones.

## Statements

### Ethical statement

The analyses conducted in this study were based on aggregated data without personal identifiers, consequently ethical review was not required.

### Use of artificial intelligence tools

None declared.

### Data availability

All model code is available on GitHub: https://doi.org/10.5281/zenodo.16783030, with a link to the GitHub repository.

## Acknowledgements

ST is funded by the NERC IAPETUS DTP (NE/S007431/1), supervised by SW, CC, BP and DB. SW, CC and DB were supported by EPSRC (EP/Y017838/1). SW and DB were supported by BBSRC-Defra (BB/X018113/1). Thanks to Santé Publique France, ARS and mosquito control operators for collecting and sharing the data.

