## Supplementary material for "Modelling and forecasting a potential chikungunya outbreak in France, 2025"


#### Contents


1. Modelling framework
2. Chikungunya transmission dynamics in small outbreak locations
3. Climate, population dynamics of *Aedes albopictus*, and chikungunya introduction dates
4. Comparison of climate between Salon-de-Provence and Toulon
5. References

### 1. Modelling framework

The model used to study the population dynamics of *Aedes albopictus* and the transmission dynamics of the chikungunya virus (CHIKV) by *Ae. albopictus* is based on a modelling framework previously developed for studying the transmission dynamics of the dengue virus by *Ae. albopictus* [1]. The model is described by a system of continuous-time delay differential equations and incorporates the trait and population dynamics of *Ae. albopictus*, including an additional phenotypic structure in the adult mosquito population that accounts for size variations (e.g., wing length). In this framework, a generic stage-structured population in class-$(i,j)$, $N_{i,j}(t)$, represents the density of individuals in life-stage $i$ and phenotypic class $j$ at time $t$, the rate of change of the population described by the system of delay-differential equations:

$$\frac{dN_{i,j}(t)}{dt} = R_{i,j}(t) - M_{i,j}(t) - D_{i,j}(t),$$

for $i = 1, \ldots, n$ and $j = 1, \ldots, m$. Here, $R_{i,j}(t)$ and $D_{i,j}(t)$ represent instantaneous recruitment and death rates for class-$(i,j)$, respectively, while $M_{i,j}(t)$ represents the instantaneous rate of maturation from the class-$(i,j)$ to the life-stage $i+1$. The recruitment rate into life-stage $i = 1$ is given by the following expression:

$$R_{1,j}(t) = \sum_{k=1}^{m} \left( w_{k,j}(\alpha(t)) \sum_{v=1}^{n} \beta_{v,k}(t) N_{v,k}(t) \right),$$

for $j = 1, \ldots, m$, where $w_{k,j}(\alpha(t))$ denotes the proportion of individuals transitioning from phenotypic class $k$ to phenotypic class $j$ at time $t$, depending on food availability $\alpha(t)$, and $\beta_{v,k}(t)$ is the birth rate of individuals in class-$(v,k)$ [2]. Maturation out of class-$(i,j)$ corresponds to the fraction of individuals that were recruited in class-$(i,j)$ $\tau_{i,j}(t)$ time units ago and survived for the stage duration $\tau_{i,j}(t)$. Formally, this is given by the relation $M_{i,j}(t) = R_{i,j}\left(t - \tau_{i,j}(t)\right) S_{i,j}(t)$, where

$$S_{i,j}(t) = \exp\left( - \int_{t-\tau_{i,j}(t)}^{t} \delta_{i,j}(s) ds \right),$$

is the probability that an individual in class-$(i,j)$ survives to progress to life-stage $i+1$ and $\delta_{i,j}(t)$ is death rate for individuals in class-$(i,j)$ and $D_{i,j}(t) = \delta_{i,j}(t) N_{i,j}(t)$. In this modelling framework, the duration of life-stage $i$, i.e., $\tau_{i,j}(t)$, also varies with time and is governed by:

$$\frac{d\tau_{i,j}(t)}{dt} = 1 - \frac{g_{i,j}(t)}{g_{i,j}\left(t - \tau_{i,j}(t)\right)},$$

where, $g_{i,j}(t)$ denotes the development rate through the class-$(i,j)$. In this modelling framework, the functions $\delta_{i,j}(t)$, $\beta_{i,j}(t)$, and $g_{i,j}(t)$ are defined by the biologically derived environment-trait relationships [1].

Finally, to analyse the transmission dynamics of viruses by mosquitoes, the mosquito population dynamics obtained through this modelling framework have been coupled with an SEIR epidemic model describing the infection status of human hosts of the virus (Supplementary Figure S1). The exposed class $(E)$ is not modelled explicitly but is instead represented through the viral incubation period within the human host. The human population is partitioned into those susceptible to infection $(H_S)$, those infected $(H_I)$, and those resistant to infection due to having recovered $(H_R)$. The size of the human population is constant, where the population density is estimated as described in the main text.

Mosquitoes are assumed to bite at a temperature-dependent rate that is inversely proportional to the length of the gonotrophic cycle. The proportion of uninfected mosquitoes of a given wing length $(A_i)$ that become infected $(I_i)$ after biting an infected human $(H_I)$ is temperature dependent. After a temperature-dependent extrinsic incubation period, an infected mosquito can bite and transmit the dengue virus to an uninfected human ($H_S$). After the intrinsic incubation period, the infected human can transmit the infection to new mosquitoes and recovers from the infection after a recovery period.

This coupled mosquito-human model has been successfully applied to study historic dengue outbreaks vectored by *Ae. albopictus* worldwide [1].

To employ this framework to investigate the transmission of CHIKV by *Ae. albopictus*. Specifically, we modelled the temperature–trait relationships (i.e., thermal reaction norms) for key model parameters: the extrinsic incubation period (EIP)—the time required for the virus to replicate and reach the mosquito's saliva following ingestion of an infected blood meal—and the probability of CHIKV transmission from *Ae. albopictus* to humans (Tegar et al. *in review*). Additionally, the temperature–trait relationship for the probability of CHIKV transmission from humans to *Ae. albopictus* was adopted from Mordecai et al. [3]. Furthermore, the intrinsic incubation period (IIP)—the duration of CHIKV incubation within humans—was assumed to have an average value of 3 days [4], while the average recovery period from CHIKV infection was taken to be 7 days [5]. We have also validated our CHIKV model against historic Italian CHIKV outbreaks; however, these results will be published elsewhere.

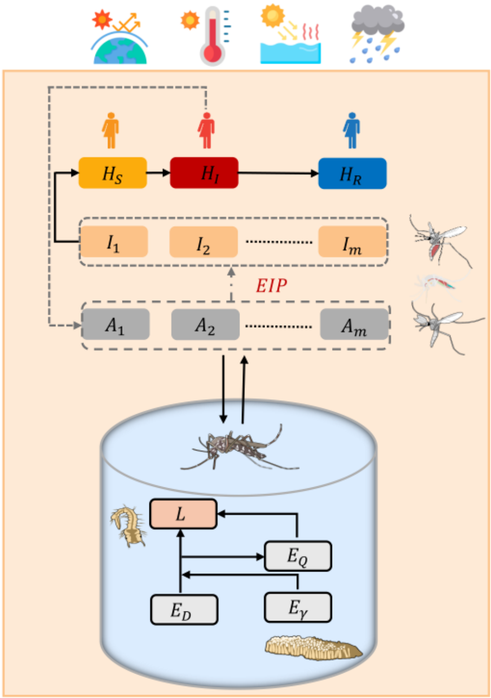


**Supplementary Figure S1.** A schematic of the climate-sensitive, stage and phenotypically structured epidemiological model. This model integrates the transmission dynamics of arboviruses by *Ae. albopictus* with the life-stage dynamics of the mosquito--eggs (diapausing: $E_D$, non-diapausing: $E_\gamma$, and quiescent: $E_Q$), larvae ($L$), and adults (suspectible $A$, infectious $I$). The adult population is further partitioned according to adult wing length, into $m$ coexisting sub-classes denoted by subscript $i$, thereby establishing phenotypic structure in the adults. The infectious mosquito sub-class comprises adult mosquitoes from class $A_i$ that have taken an infected blood meal and then gone on to complete the extrinsic incubation period (EIP), defined as the average time required for the virus to incubate within the mosquito. The human population is represented by three compartments: susceptible ($H_S$), infected ($H_I$), and recovered ($H_R$).

#### 2. Chikungunya transmission dynamics in small outbreak locations

This section presents the predicted CHIKV transmission dynamics for locations in France for which the model simulations predicted small chikungunya outbreaks: Lipsheim (Bas-Rhin, Grand Est), Saint-Chamond (Loire, Auvergne-Rhône-Alpes), Claix (Isère, Auvergne-Rhône-Alpes), Montoison (Drôme, Auvergne-Rhône-Alpes), and Grosseto-Prugna (Corse du Sud, Corse).

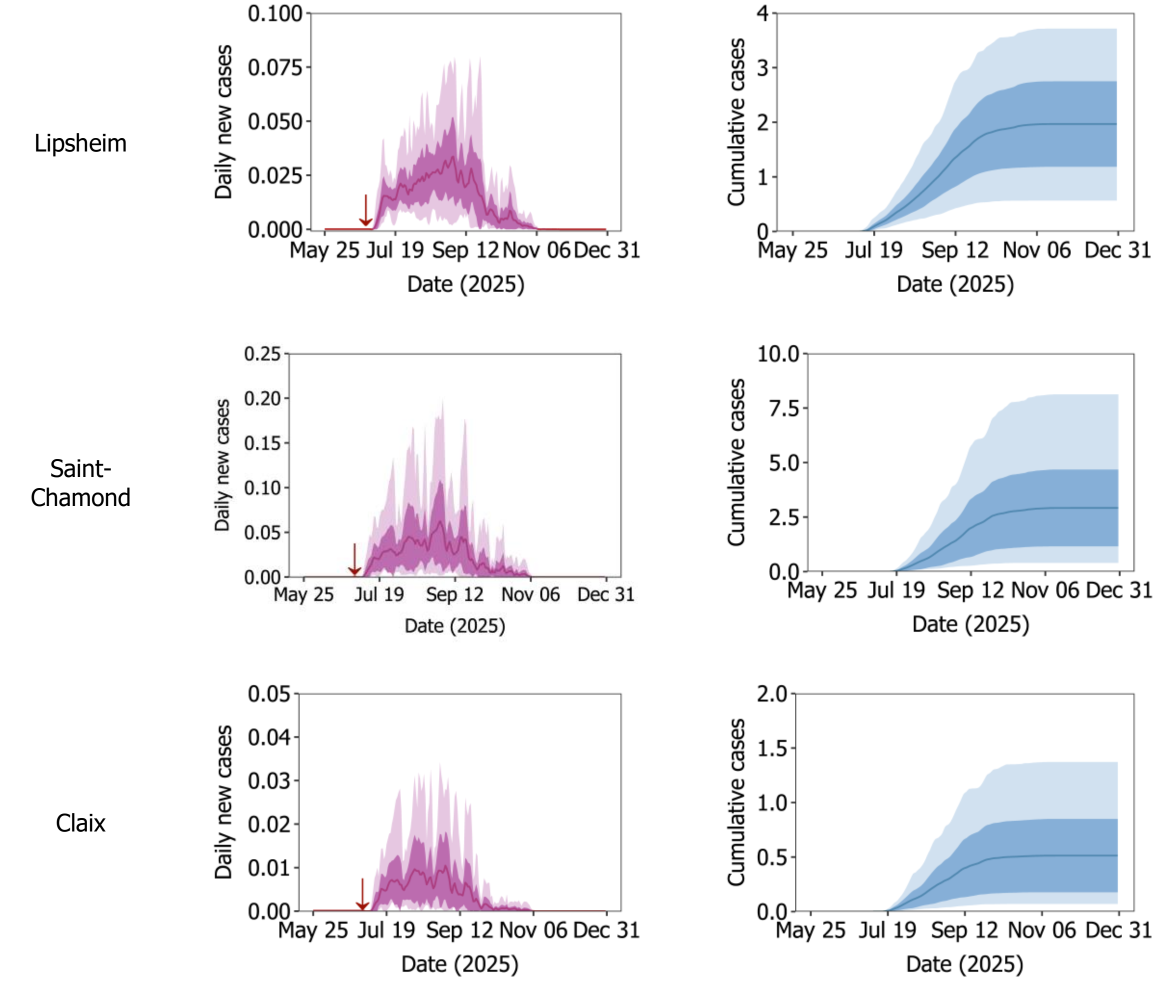


**Supplementary Figure S2 (part 1).** CHIKV transmission dynamics for locations in France where smaller outbreaks are predicted. For each location shown in the panels above, the red plots represent the daily number of cases. The blue plots show the cumulative number of cases. In all plots, the solid lines represent the mean values, the dark ribbons indicate the 95% confidence intervals (CIs), and the light-coloured bands correspond to the absolute maximum and minimum values. The introduction dates, indicated by red downward arrows in the plots, correspond to the first reported symptomatic case in each location (Table 1). The variability in the projections reflects the estimated stochasticity in climate variables—temperature, precipitation, and evaporation—for the remainder of the year 2025, see SI3 for details.

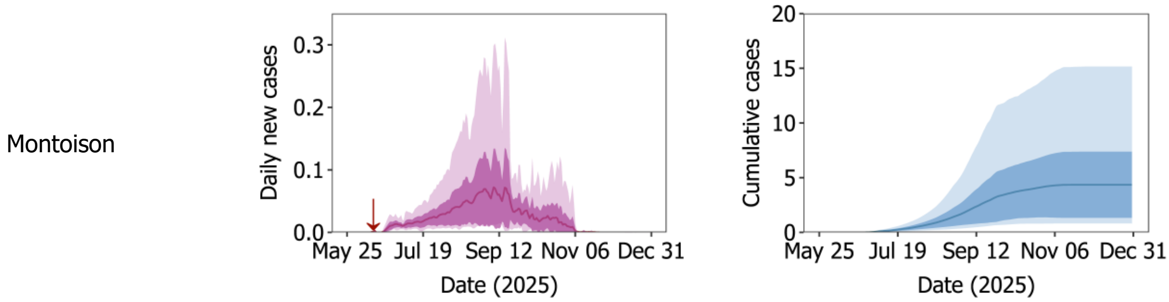

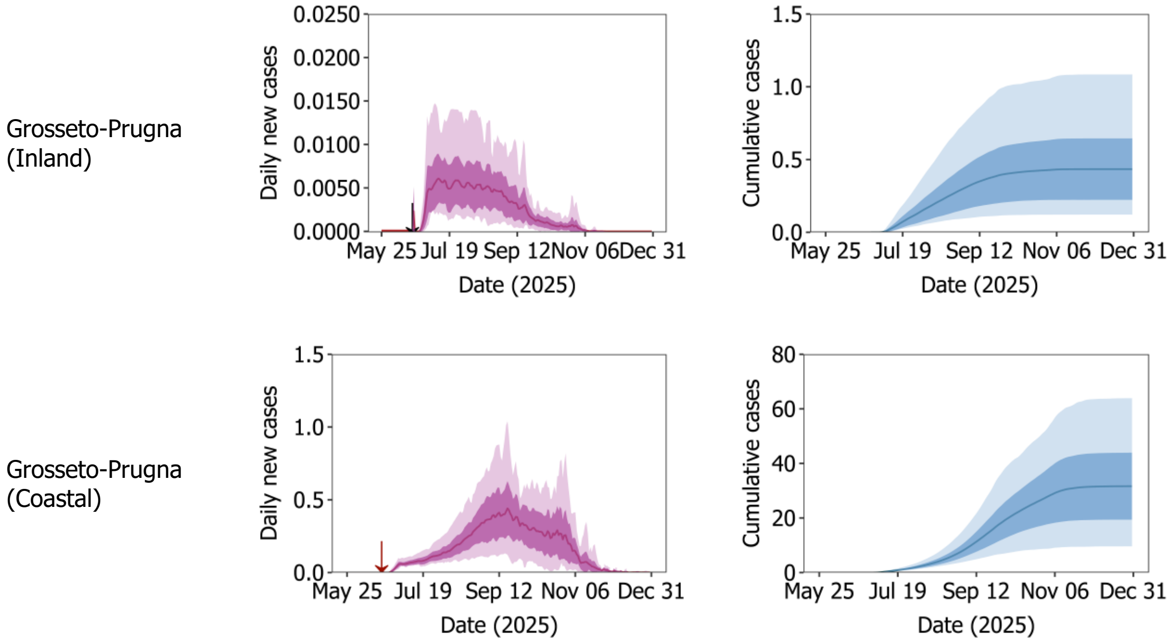


**Supplementary Figure S2 (part 2).** CHIKV transmission dynamics for locations in France where smaller outbreaks could occur. For each location shown in the panels above, the red plots represent the daily number of cases. The blue plots show the cumulative number of cases. In all plots, the solid lines represent the mean values, the dark ribbons indicate the 95% confidence intervals (CIs), and the light-coloured bands correspond to the absolute maximum and minimum values. The introduction dates, indicated by red downward arrows in the plots, correspond to the first reported symptomatic index case in each location (Table 1). The variability in the projections reflects the estimated stochasticity in climate variables—temperature, precipitation, and evaporation—for the remainder of the year 2025, see SI3 for details.

#### 3. Climate, population dynamics of *Aedes albopictus*, and chikungunya introduction dates

This section presents the climate scenarios (temperature and rainfall) considered for projecting the model forward in time beyond June 13 2025, when recorded climate date ends. The climate scenarios are based on data from June 13 to December 31 from the previous ten years (2015–2024). Also presented are model estimated adult *Ae. albopictus* population dynamics, oviposition activities, and the total cumulative number of chikungunya cases at the end of epidemic (i.e., the final epidemic size) for each outbreak location reported up to 16 July 2025 [6]: La Crau (Var, Provence-Alpes-Côte d'Azur), Prades-le-Lez (Hérault, Occitanie), Salon-de-Provence (Bouches-du-Rhône, Provence-Alpes-Côte d'Azur), Grosseto-Prugna (Corse-du-Sud, Corse), Montoison (Drôme, Auvergne-Rhône-Alpes), Bernis (Gard, Occitanie), Lipsheim (Bas-Rhin, Grand Est), Claix (Isère, Auvergne-Rhône-Alpes), Fréjus (Var, Provence-Alpes-Côte d'Azur), Saint-Brès/Castries (Hérault, Occitanie), Toulon (Var, Provence-Alpes-Côte d'Azur, and Saint-Chamond (Loire, Auvergne-Rhône-Alpes).

Each of the panel plots in Figures S3-S16 includes: (A) daily temperature, (B) daily total precipitation, (C) dynamics of adult and egg stages of *Ae. albopictus*, and (D) a boxplot of the final outbreak sizes (total cumulative chikungunya cases at the end of the season) for each introduction date, showing variation across ten climate scenarios from 2015 to 2024. Dots represent outliers in some studied cities, indicating exceptionally higher final sizes for specific climate scenarios, mostly corresponding to years warmer than the average annual temperature (e.g., 2022 for La Crau; Supplementary Figure S9). In subplots (A–C), the solid lines represent mean values, the dark ribbons indicate 95% confidence intervals (CIs), and the light-coloured bands correspond to the absolute maximum and minimum values.

#### Bernis

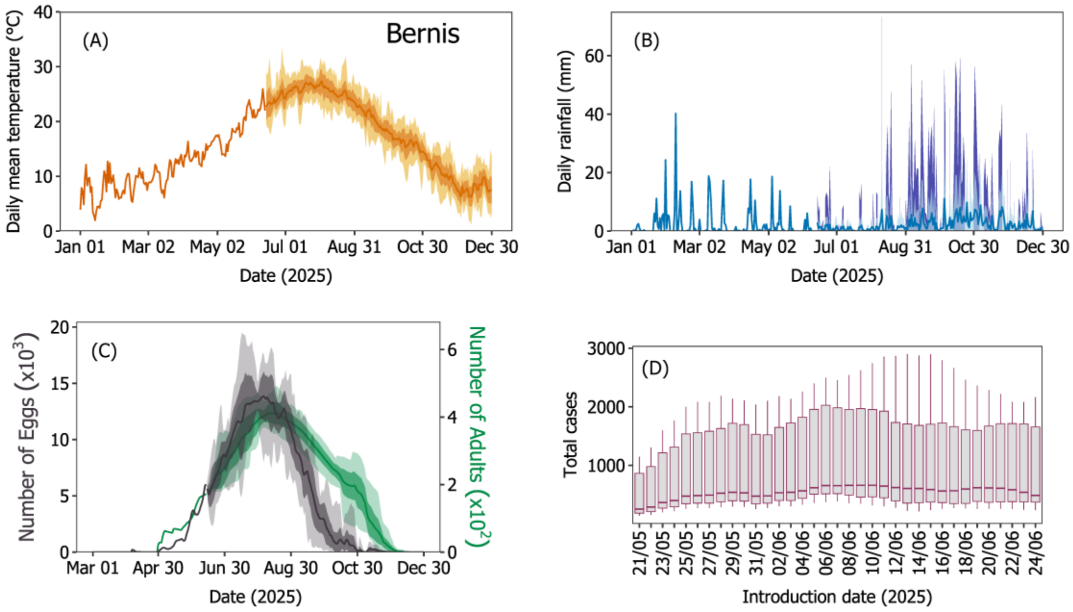


**Supplementary Figure S3.** Temperature, rainfall, adult *Ae. albopictus* dynamics with oviposition activity, and total predicted cases estimated for introduction dates spanning 3 weeks before to 2 weeks after the first reported symptomatic index case in Bernis.

#### Castries

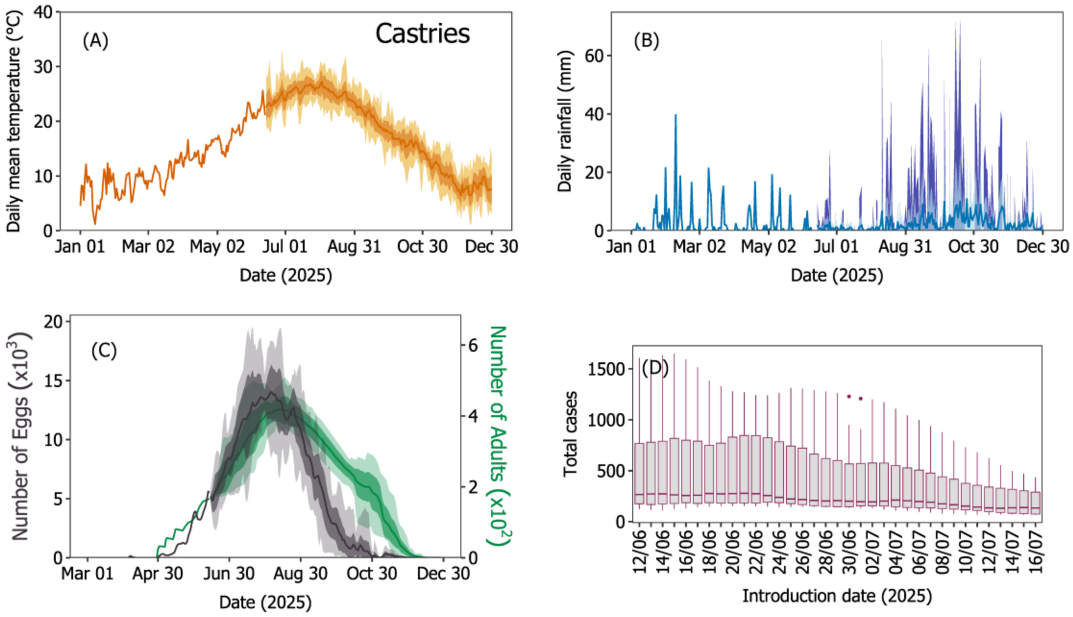


**Supplementary Figure S4.** Temperature, rainfall, adult *Ae. albopictus* dynamics with oviposition activity, and total predicted cases estimated for introduction dates spanning 3 weeks before to 2 weeks after the first reported symptomatic index case in Castries. Dots represent outliers.

#### Claix

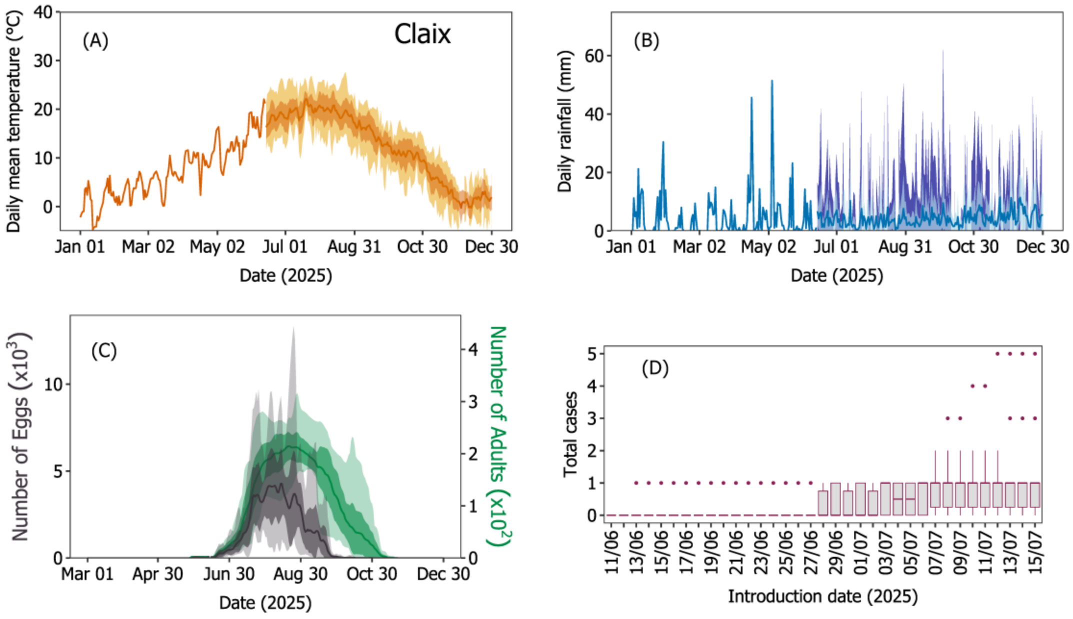


**Supplementary Figure S5.** Temperature, rainfall, adult *Ae. albopictus* dynamics with oviposition activity, and total predicted cases estimated for introduction dates spanning 3 weeks before to 2 weeks after the first reported symptomatic index case in Claix. Dots represent outliers.

#### Fréjus

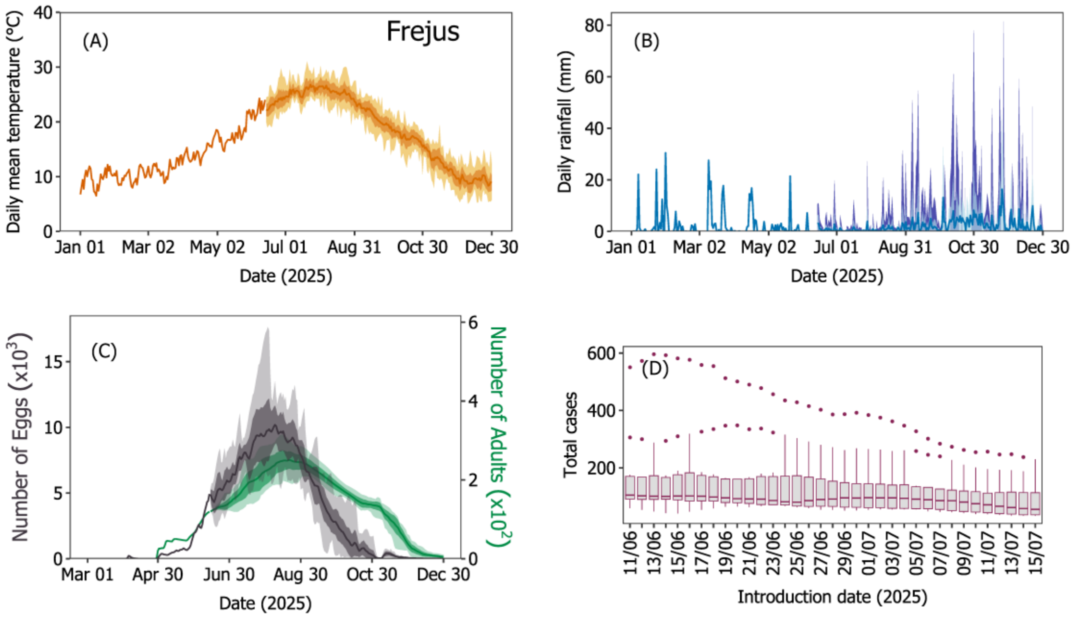


**Supplementary Figure S6.** Temperature, rainfall, adult *Ae. albopictus* dynamics with oviposition activity, and total predicted cases estimated for introduction dates spanning 3 weeks before to 2 weeks after the first reported symptomatic index case in Fréjus. Dots represent outliers.

#### Grosseto-Prugna (Coastal)

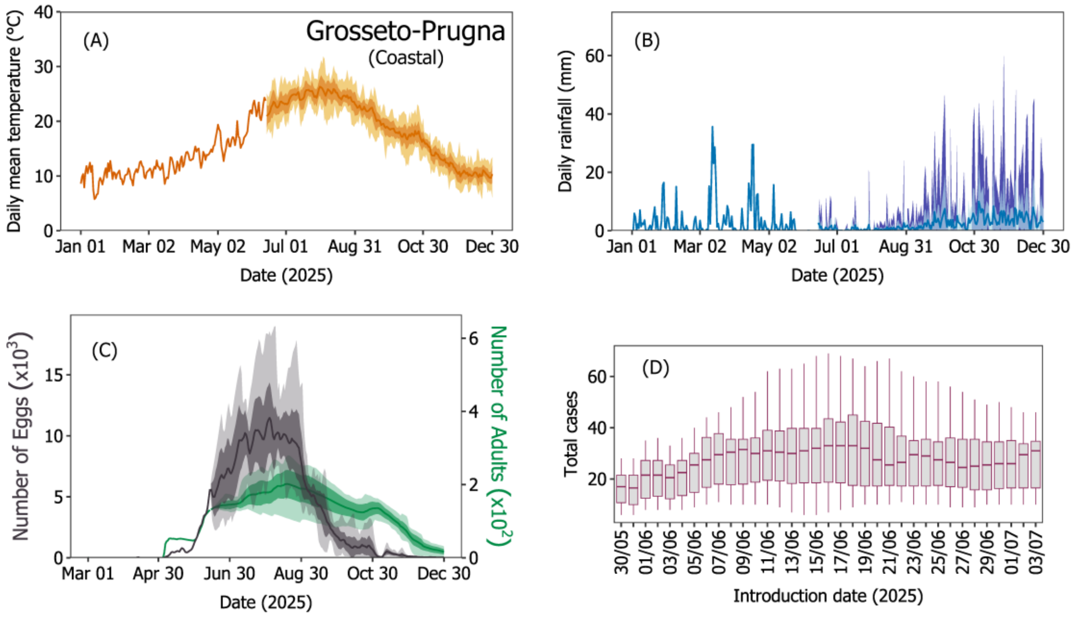


**Supplementary Figure S7.** Temperature, rainfall, adult *Ae. albopictus* dynamics with oviposition activity, and total predicted cases estimated for introduction dates spanning 3 weeks before to 2 weeks after the first reported symptomatic index case in Grosseto-Prugna (coastal).

#### Grosseto-Prugna (inland)

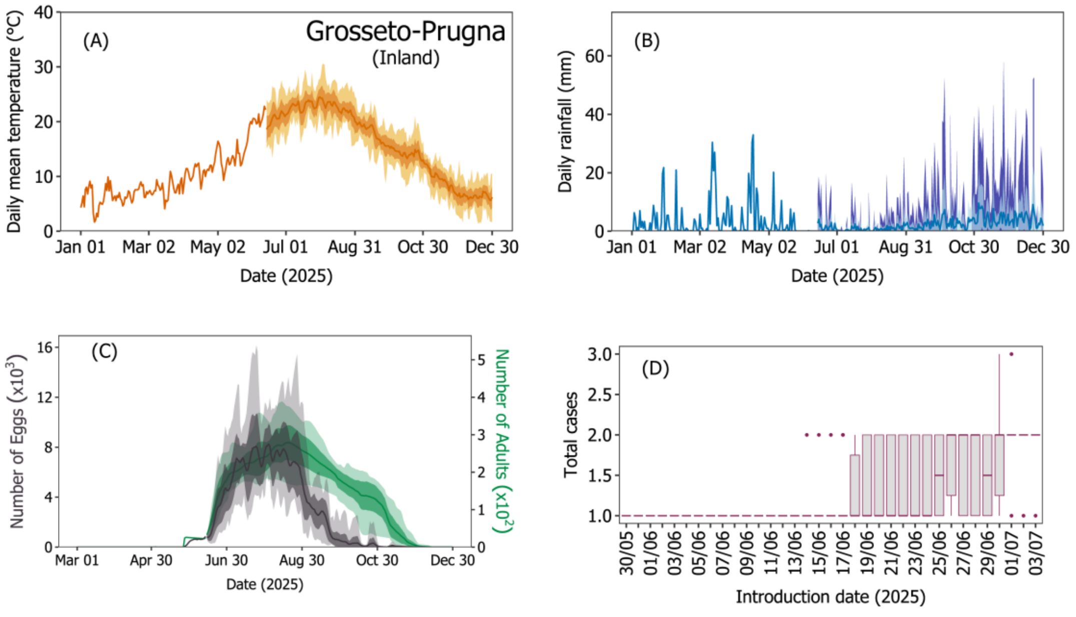


**Supplementary Figure S8.** Temperature, rainfall, adult *Ae. albopictus* dynamics with oviposition activity, and total predicted cases estimated for introduction dates spanning 3 weeks before to 2 weeks after the first reported symptomatic index case in Grosseto-Prugna (inland). Dots represent outliers.

#### La Crau

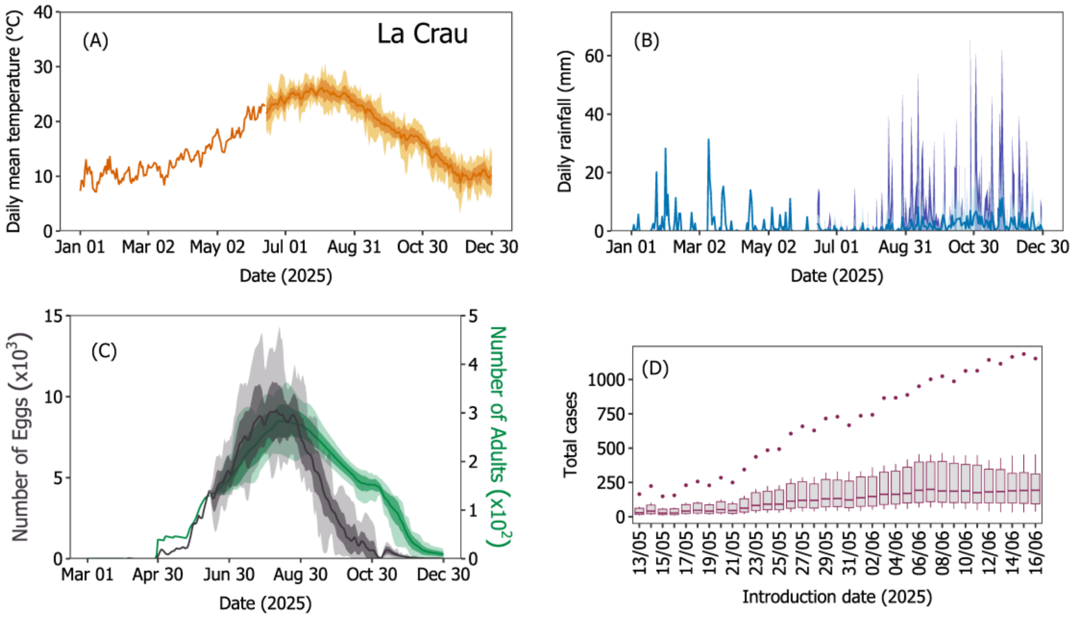


**Supplementary Figure S9.** Temperature, rainfall, adult *Ae. albopictus* dynamics with oviposition activity, and total predicted cases estimated for introduction dates spanning 3 weeks before to 2 weeks after the first reported symptomatic index case in La Crau. Dots represent outliers.

#### Lipsheim

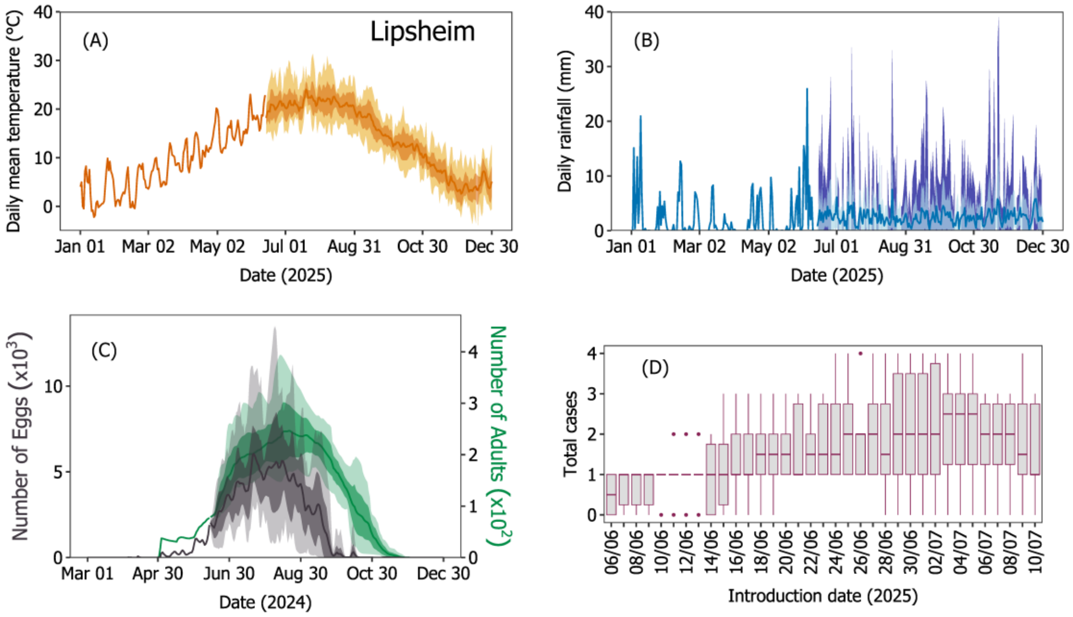


**Supplementary Figure S10.** Temperature, rainfall, adult *Ae. albopictus* dynamics with oviposition activity, and total predicted cases estimated for introduction dates spanning 3 weeks before to 2 weeks after the first reported symptomatic index case in Lipsheim. Dots represent outliers.

#### Montoison

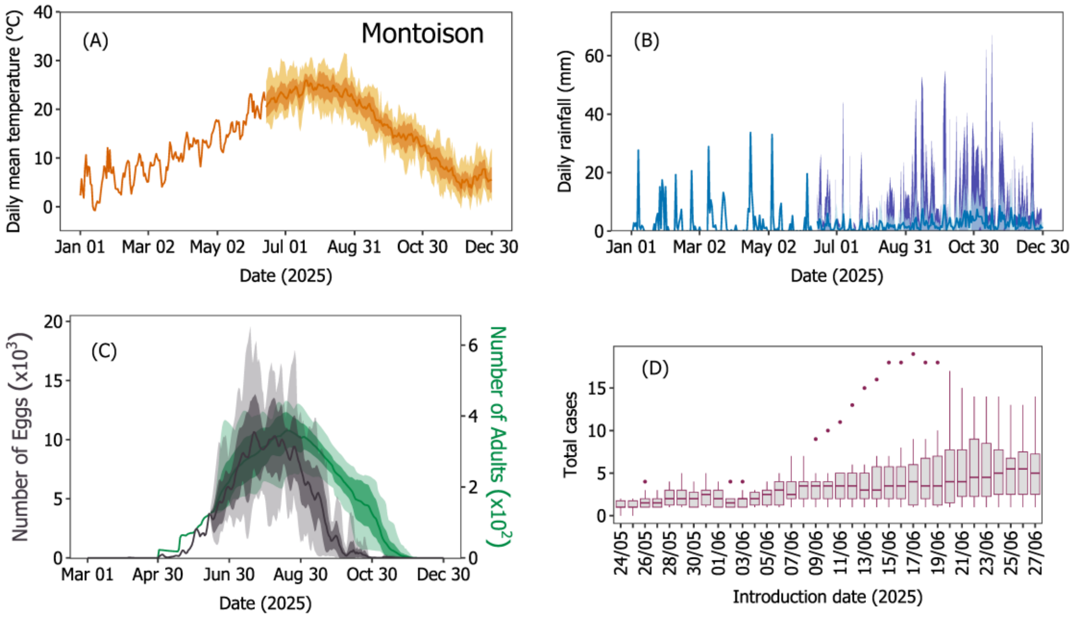


**Supplementary Figure S11.** Temperature, rainfall, adult *Ae. albopictus* dynamics with oviposition activity, and total predicted cases estimated for introduction dates spanning 3 weeks before to 2 weeks after the first reported symptomatic index case in Montoison. Dots represent outliers.

#### Prades-le-Lez

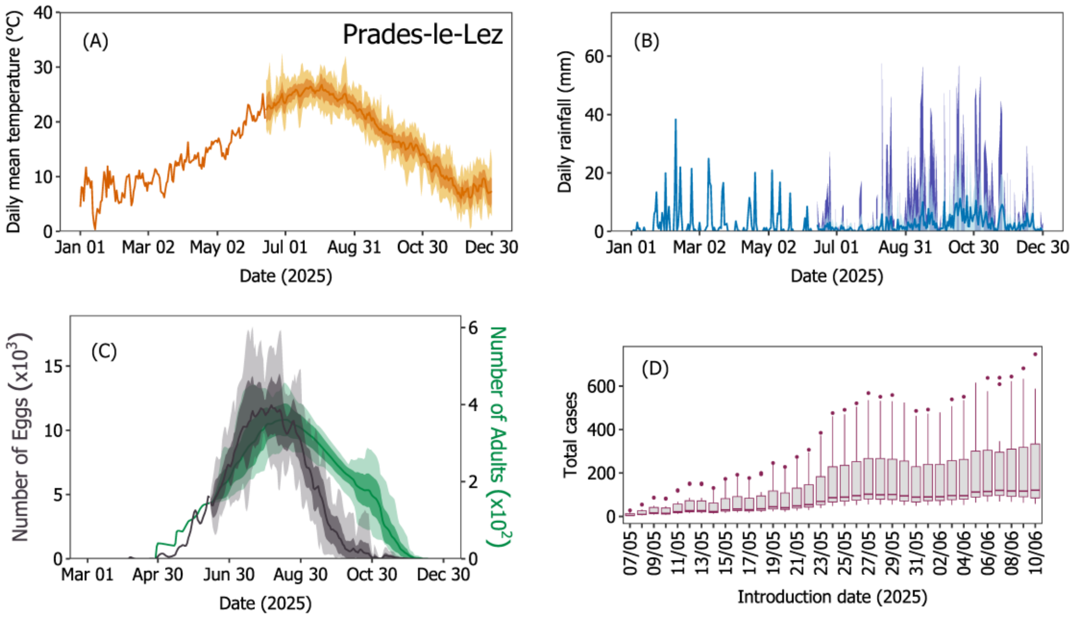


**Supplementary Figure S12.** Temperature, rainfall, adult *Ae. albopictus* dynamics with oviposition activity, and total predicted cases estimated for introduction dates spanning 3 weeks before to 2 weeks after the first reported symptomatic index case in Prades-le-Lez. Dots represent outliers.

#### Salon-de-Provence

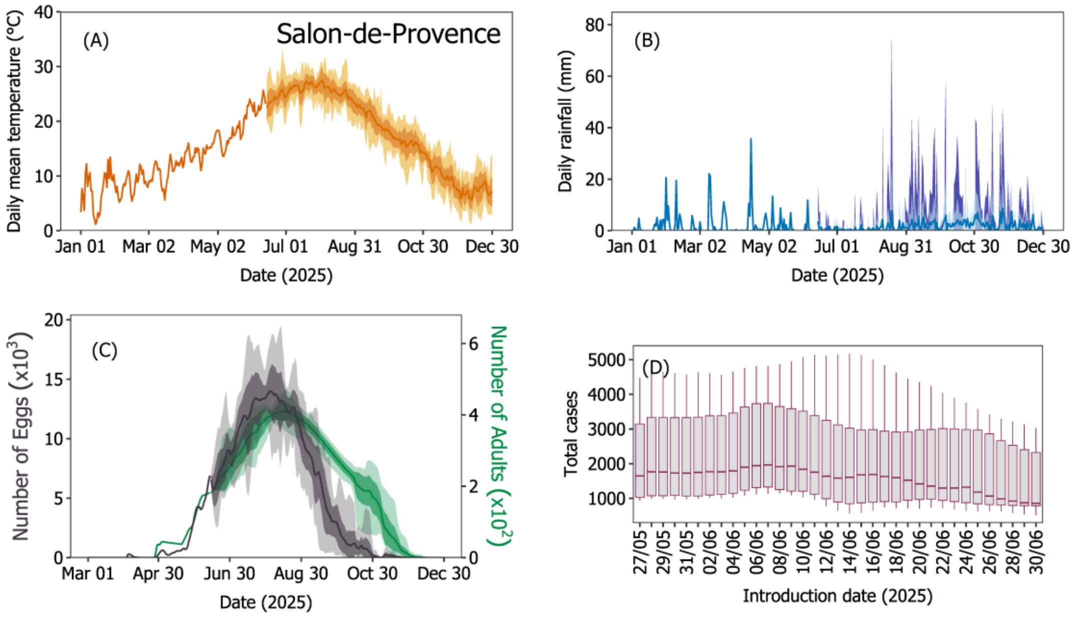


**Supplementary Figure S13.** Temperature, rainfall, adult *Ae. albopictus* dynamics with oviposition activity, and total predicted cases estimated for introduction dates spanning 3 weeks before to 2 weeks after the first reported symptomatic index case in Salon-de-Provence.

#### Saint Bres

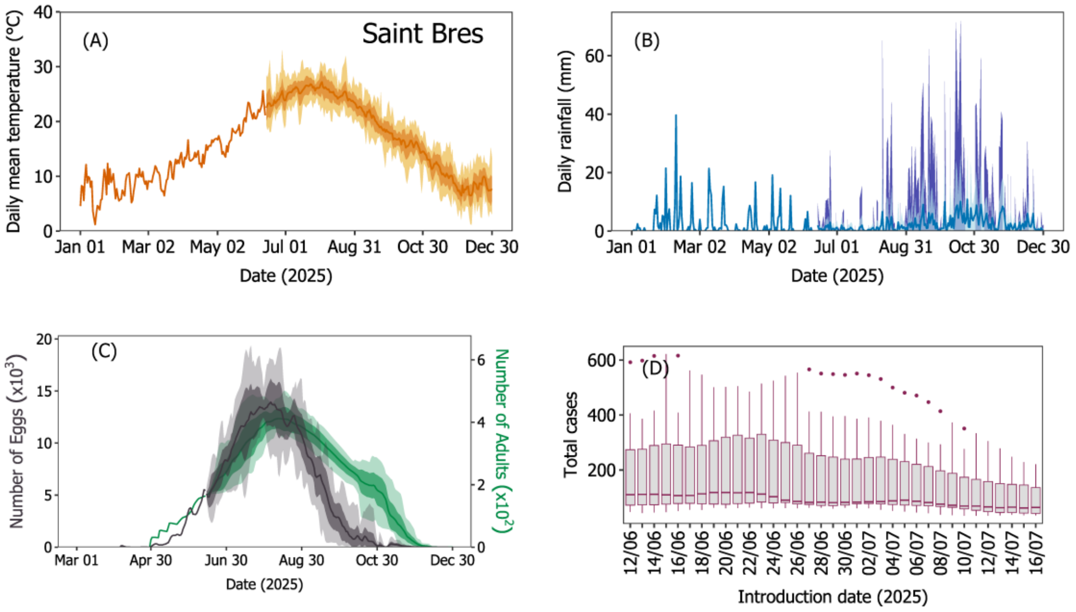


**Supplementary Figure S14.** Temperature, rainfall, adult *Ae. albopictus* dynamics with oviposition activity, and total predicted cases estimated for introduction dates spanning 3 weeks before to 2 weeks after the first reported symptomatic index case in Saint Bres. Dots represent outliers.

#### Saint-Chamond

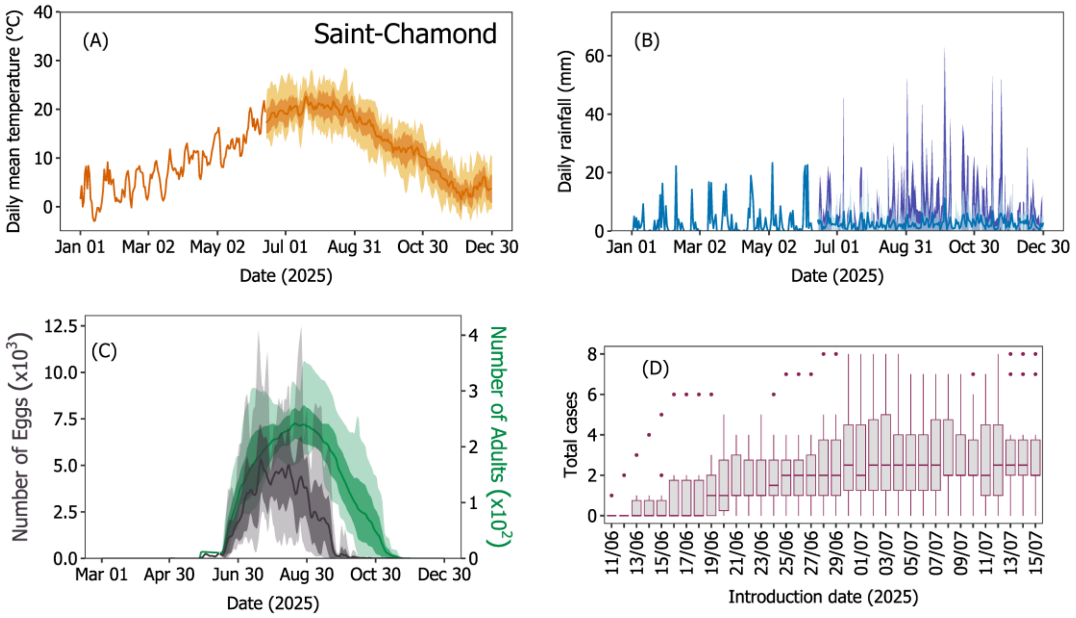


**Supplementary Figure S15.** Temperature, rainfall, adult *Ae. albopictus* dynamics with oviposition activity, and total predicted cases estimated for introduction dates spanning 3 weeks before to 2 weeks after the first reported symptomatic index case in Saint-Chamond. Dots represent outliers.

#### Toulon

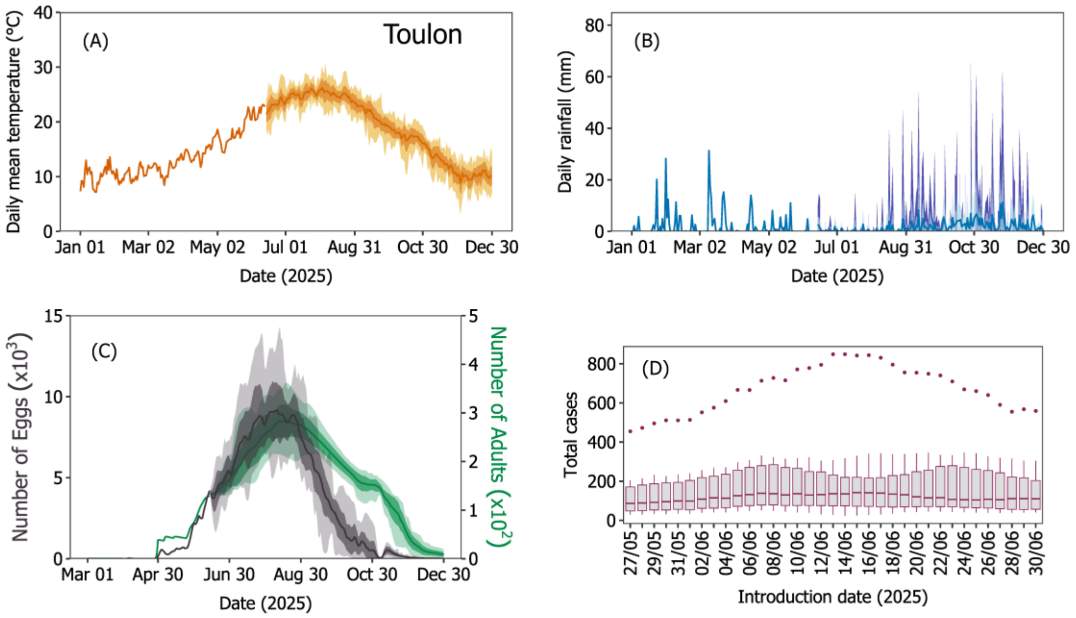


**Supplementary Figure S16.** Temperature, rainfall, adult *Ae. albopictus* dynamics with oviposition activity, and total predicted cases estimated for introduction dates spanning 3 weeks before to 2 weeks after the first reported symptomatic index case in Toulon. Dots represent outliers.

#### 4. Comparison of the climate between Salon-de-Provence and Toulon

For the simulations of Salon-de-Provence and Toulon, we identified four geographical blocks with higher human density, each measuring 2 km by 2 km, covering a total area of 16 km² in each city. We extracted climate data from the ERA5 Land reanalysis dataset [7] for each of these four blocks and ran the model separately for each block. Finally, we averaged the results across the four blocks to represent the climate, mosquito population, and transmission dynamics over the entire 16 km² area for each city. Notably, the months corresponding to the active life cycle of *Ae. albopictus* and arbovirus transmission (e.g., April–August) appear to be more climatically favourable in Salon-de-Provence than in Toulon (Supplementary Figure S17). Climate data for the year 2025 was available only up to June 13, 2025. Beyond this date, historical climate data from 2015 to 2024 were used to represent the remainder of 2025, resulting in 10 climate scenarios based on each year from 2015 to 2024 for the period after June 13, 2025.

This subtle difference—particularly during the early part of the season (April–August) in a year—may influence several key eco-epidemiological traits of *Ae. albopictus*, including speedy development during aquatic life stages, increased adult locomotor activity, shortened gonotrophic cycles, increased biting activity, and reduced viral incubation periods. Collectively, these factors may contribute to higher *Ae. albopictus* population densities, increased biting rates, and a greater potential for chikungunya virus transmission in Salon-de-Provence compared to Toulon (see Supplementary Figure S13, S15 & S17).

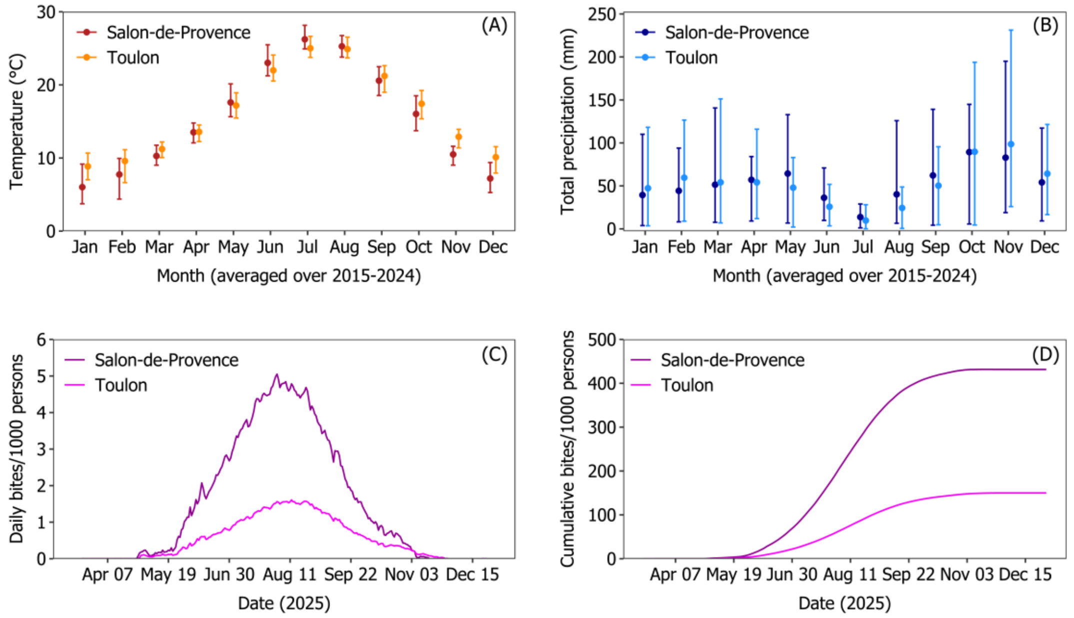


**Supplementary Figure S17.** Comparison of monthly temperature, precipitation, and number of *Ae. albopictus* bites per 1,000 persons between Salon-de-Provence and Toulon. Climate variables—(A) average monthly temperature and (B) total monthly precipitation—are presented as ten-year means (2015–2024) to illustrate typical seasonal patterns. Panels (C) and (D) show, respectively, the daily number of *Ae. albopictus* bites per 1,000 persons and the cumulative number of bites per 1,000 persons over the course of the season.
